# Unsupervised clustering reveals phenotypes of AKI in ICU Covid19 patients

**DOI:** 10.1101/2022.03.11.22272259

**Authors:** Frederic Sangla, Elisa Marchi, Benjamin Assouline, Christophe Leterrier, Sebastian Sgardello, Jérôme Pugin, Gilles Criton, David Legouis

## Abstract

**Background:** Acute Kidney Injury (AKI) is a very frequent condition, occurring in about one in three patients admitted to an intensive care unit (ICU). AKI is a syndrome defined as a sudden decrease in glomerular filtration rate. However, this unified definition does not reflect the various mechanisms involved in AKI pathophysiology, each with its own characteristics and sensitivity to therapy. In this study, we aimed at developing an innovative machine learning based method able to subphenotype AKI according to its pattern of risk factors.

**Methods:** We adopted a three-step pipeline of analyses. Firstly, we looked for factors associated with AKI using a generalized additive model. Secondly, we calculated the importance of each identified AKI related factor in the estimated AKI risk to find the main risk factor for AKI, at the single patient level. Lastly, we clusterized AKI patients according to their profile of risk factors and compared the clinical characteristics and outcome of every cluster. We applied this method to a cohort of severe Covid19 patients hospitalized in the ICU of Geneva University Hospitals.

**Results:** Among the 250 patients analyzed, we found ten factors associated with AKI development. Using the individual expression of these factors, we identified three groups of AKI patients, based on the use of Lopinavir/Ritonavir, a prior history of diabetes mellitus and baseline eGFR and ventilation. The three clusters expressed distinct characteristic in terms of AKI severity and recovery, metabolic patterns and ICU mortality.

**Conclusion:** We propose here a new method to phenotype AKI patients according to their most important individual risk factors for AKI development. When applied to an ICU cohort of Covid19 patients, we were able to differentiate three groups of patients. Each expressed specific AKI characteristics and outcomes, which probably reflects a distinct pathophysiology.

## Background

Acute Kidney Injury (AKI) is a common condition in the critical care setting[1,2]. Despite decades of research, AKI is still associated with high mortality and morbidity, even when renal function is substituted by Renal Replacement Therapy (RRT)[3–6].

AKI is defined as a sudden decrease in glomerular filtration rate, demonstrated by an increase in serum creatinine[7]. This unified definition has resulted in an improved recognition of AKI and has simplified research, healthcare management as well as comparisons across cohorts and different centers. However, AKI is not a single clinical entity but an overarching clinical syndrome. Therefore, the definition of AKI encompasses many underlying conditions and etiologies. Additionally, the high degree of heterogeneity of the Intensive Care Unit (ICU) population including patients with different risk profile adds further complexity when considering AKI outcomes[8]. In this respect, recognizing meaningful subgroups of AKI patients may provide a deeper insight in AKI pathophysiology and may also be helpful in identifying groups with differing prognoses and sensitivity to therapy [9].

From a data-driven perspective, patient sub-phenotyping is essentially a clustering problem[10,11]. Clustering algorithms are a type of unsupervised machine learning algorithm where no labels are known a priori but get assigned based on inherent similarities between points. A critical step in clusterisation is the representation of the data i.e., the construction of the dataset on which we want to apply clusterisation. Previous studies on AKI sub-phenotyping have defined patients according to diagnostic codes[12], trajectories of serum creatinine[13], patterns of AKI reversal[14] or clinical and biological data recorded at ICU admission[15] or during AKI[16,16,17]. However, those strategies do not allow for the formulation of any hypothesis based on the pathophysiological mechanisms involved in different AKI phenotypes. In addition, the high number of features used to classify patients makes it difficult to recognize them at the bedside, in current practice.

In this study, we aimed at developing an innovative pipeline of analyses in order to identify in an unsupervised manner, distinct phenotypes of AKI in ICU Covid19 patients, based on their pattern of AKI associated factors.

## Methods

### Patient inclusion

During the study period from March to December 2020, all Covid-19 patients admitted to the adult intensive care unit of the Geneva University Hospitals were screened. Patients were included if they were older than 18 years of age, did not develop AKI prior to ICU admission and were not on chronic dialysis. The study was conducted according to the guidelines of the Declaration of Helsinki, and approved by the ethical committee for human studies of Geneva, Switzerland (CCER 2020-00917, Commission Cantonale d’Ethique de la Recherche).

### Definitions

AKI was defined according to the KDIGO criteria[7], i.e. a 1.5 fold or more increase in baseline serum creatinine levels within 7 days or, an absolute increase higher than 26.4 μmol/L within 48 hours. Baseline serum creatinine levels were determined as the first serum creatinine level recorded following hospital admission.

If serum creatinine levels at ICU discharge were equal to or were below baseline serum creatinine levels, patients were considered as having recovered from AKI within the ICU. This outcome was calculated only in patients discharged alive from ICU.

Mid-term recovery was defined as a decrease in 50% of serum creatinine compared to the maximal level reached during ICU stay.

### Data collection

For each patient, the following variables were recorded: demographical data (sex, age, body mass index, height and weight), prior history of hypertension, diabetes, Chronic Obstructive Pulmonary Disease (COPD), hypercholesterolemia, smoking, cardiomyopathy and heart failure, cerebrovascular disease, malignancy, chronic kidney disease (defined as a history of chronic renal disease in the patient’s medical records), chronic use of Non-Steroidal Anti Inflammatory Drugs (NSAIDs), renin angiotensin aldosterone system inhibitors or steroids. Upon ICU admission, we recorded biological data (prothrombin ratio, procalcitonin, C-reactive protein, d-dimer, white blood cells, lymphocytes, neutrophils, thrombocytes, lactate, bilirubin, alanine transaminase (TGP), aspartate transaminase (TGO), troponin levels, serum creatinine and eGFR), severity scores (APACHE, SAPS, SOFA) and the FiO2. Once patients were intubated, we recorded the initial respiratory parameters (PaO2/FiO2 ratio, PEEP and plateau pressure levels, compliance, tidal volume, duration from symptoms or hospitalisation to intubation, respiratory rate before intubation). Finally, we screened the following variables for the entire ICU stay: the need for invasive mechanical ventilation, Neuro Muscular Blocking Agents (NMBA), Extra Corporeal Membrane Oxygenation (ECMO), norepinephrine, antibiotics and their total duration, the need for prone positioning and the number of prone sessions, the use of Lopinavir/Ritonavir (LPV/r), hydroxychloroquine, azithromycin, remdesivir, anakinra, dexamethasone and inhaled nitric oxide. At the renal level, we collected all the serum creatinine recorded during the hospital stay, as well as the need for renal replacement therapy. We also recorded the time between symptoms and admission to hospital, ICU and intubation, the duration between hospital and ICU admission and intubation. Glucose and lactate levels measured during the ICU stay were also collected.

### Statistical analysis

Baseline characteristics were expressed as mean (standard deviation) and median (25-75^th^ percentiles) or absolute and relative (%) frequency if categorical. They were compared using a Mann Whitney or Chi-square tests depending on their class. A p-value of less than 0.05 was considered significant

### Data pre-processing

Numerical data were centred, scaled and underwent Yeo-Johnson transformation. Missing data were further imputed using bagged tree imputation [18]. This step was completed using the *caret* package.

### Identification of factors associated with AKI development

To identify factors associated with AKI development, we first fitted for each recorded variable, a univariable logistic regression modelling the logit of AKI. Variables displaying a p-value below 0.2 were first considered for the multivariable analyses. This was conducted using a generalized additive model to allow nonlinear relationships that were fitted using thin plate regression splines from the *mgcv* package. Variable selection was further performed using a supervised stepwise approach, as previously described, to only keep predictors with a p-value lower than 0.05 [19,20]. Validation of the nonlinear fitting was achieved by building a second generalized additive model. Instead of regression splines, local regression was used by locally estimating scatterplot smoothing curve fitting, as supported by the *gam* package,. The two nonlinear fits were further visually compared by displaying the partial dependence plots of each model.

Discrimination and calibration of the final model were visually assessed through the receiver operating characteristic (ROC) curve and a calibration plot and numerically by the calculation of the area under the ROC curve and the Hosmer-Lemeshow test.

### ML based validation of the feature selection

To validate the supervised variable selection performed in the generalized additive model, we used an unsupervised approach using 4 machine learning methods that integrate native automated feature selection: multivariate adaptative regression spline (MARS), multistep adaptative MCPnet, lasso regression and regularized random forest (RRF). These four algorithms were applied on the whole dataset, tuning the hyperparameters by using 5 repetitions of 10 cross-validations. The out-of-bag area under the ROC curve was recorded at each time.

### SHAP values

Shapley Additive Explanation values were calculated using the *shapr* package using an empirical approach.

### Clusterisation

A Matrix of SHAP values was used as an input for Uniform Manifold Approximation and Projection (UMAP), using a Euclidean metric, a minimal distance of 0.1 and 15 neighbors. Patients projected on this UMAP were further clusterized using an unsupervised method, the Density-Based Spatial Clustering of Application with Noise (DBSCAN) algorithm, through the *dbscan* package. The radius of the epsilon neighborhood was set to 1. The resulting clusterisation was further validated by linear support vector machines (SVM), applied to each cluster against the others. For this purpose, the SHAP matrix was first split in a train and a test dataset using a 0.8:0.2 ratio. Five SVM were first trained on test dataset, in order to tune their hyperparameters to maximize the area under the ROC curve. 3 repetitions of 10 cross-validations were used, with each SVM predicting one cluster against the 4 others. The optimal SVM models were further applied on the 2000-fold bootstrapped test datasets.

### Metabolic pattern

For each patient, we calculated the relative time spent in one of the five metabolic patterns previously described [21,22], *i*.*e*. the total duration spent in each of the five profiles divided by the total duration of ICU stay.

### Clinical outcomes

For each of the three clusters, we used a Naïve Bayes algorithm to calculate the a posteriori probability of each outcome (*i*.*e*. ICU mortality, RRT weaning, renal recovery and relative time spent in each metabolic pattern) within each cluster. We used resampling by bootstrap (n=2000) to estimate confidence intervals and p-values.

## Results

### Cohort Description

From March to December 2020, 256 Covid-19 patients were admitted to ICU of the Geneva University Hospitals. Among them, 6 were not included for the following reasons: one patient developed AKI prior ICU admission and 5 patients were on chronic dialysis. A total of 250 patients were analyzed, of which 104 (42%) experienced AKI during their ICU stay. Most of them developed KDIGO1 AKI (68%) while 14 (13%) received Renal Replacement Therapy (RRT). Compared to those who did not develop AKI, AKI patients more frequently reported a history of diabetes, hypertension and hypercholesterolemia. They had a lower estimated Glomerular Filtration Rate (eGFR) at hospital entry, were older and mostly male. Furthermore, they had higher APACHE and SOFA scores as well as troponin, C reactive protein and procalcitonin levels but lower lactate levels at ICU admission. During their ICU stay, AKI patients were more likely to receive norepinephrine, Lopinavir/Ritonavir (LPV/r), hydroxychloroquine, azithromycin and neuromuscular blocking agents (NMBA), but not dexamethasone. Finally, AKI patients more frequently required invasive mechanical ventilation and prone positioning, received higher tidal volumes, spent more time on mechanical ventilation and had longer ICU and hospital lengths of stay. Time between onset of symptoms and intubation was longer. However, mortality was not different between AKI and non-AKI patients. Table 1 shows all the compared characteristics between these two groups.

**Table 1.** Baseline Characteristics: Data are presented as mean (percentage) or as median (interquartile range). BMI Body Mass Index; eGFR estimated glomerular filtration rate; FiO2 = Oxygen inspirited fraction; SAPS II Simplified Acute Physiological Score; APACHE II Acute Physiology and Chronic Health Evaluation; SOFA Sepsis-Related Organ Failure Assessment; KDIGO Kidney Disease: Improving Global Outcomes; RRT Continuous Renal Replacement Therapy; LPV/r Lopinavir/Ritonavir; NSAID Non-Steroidal Anti Inflammatory Drug; NMBA Neuro Muscular Blocking Agents; ECMO Extra Corporeal Membrane Oxygenation; P/F PaO2/FiO2 ratio; ICU Intensive Care Unit; AKI Acute Kidney Injury; LOS Length of Stay

|  | No-AKI (N=146) | AKI (N=104) | Total (N=250) | pvalue |
| --- | --- | --- | --- | --- |
| **Patients' characteristics** |  |  |  |  |
| Age (years), | 63.0 (55.0, 73.0) | 67.0 (59.0, 74.0) | 65.5 (57.0, 74.0) | 0.021 |
| Sex, male, n (%) | 101 (69.2%) | 88 (84.6%) | 189 (75.6%) | 0.007 |
| Weight (cm) | 81.3 (70.0, 95.0) | 85.0 (73.8, 100.2) | 83.8 (71.0, 98.0) | 0.052 |
| BMI (mkg/m<sup>2</sup>) | 27.8 (24.6, 31.3) | 28.5 (25.4, 32.8) | 27.9 (24.9, 32.3) | 0.080 |
| Hypertension, n (%) | 56 (38.4%) | 58 (55.8%) | 114 (45.6%) | 0.007 |
| Diabetes, n (%) | 36 (24.7%) | 43 (41.3%) | 79 (31.6%) | 0.006 |
| Chronic kidney disease, n (%) | 6 (4.1%) | 14 (13.5%) | 20 (8.0%) | 0.009 |
| **During ICU stay** |  |  |  |  |
| eGFR at hospital admission | 67.7 (51.9, 92.1) | 50.1 (34.8, 65.9) | 59.5 (43.5, 83.5) | < 0.001 |
| FiO2 at ICU admission (%) | 60.0 (50.0, 80.0) | 63.0 (50.0, 77.8) | 60.0 (50.0, 80.0) | 0.907 |
| SAPS II score | 51.5 (35.0, 64.0) | 55.0 (41.8, 67.2) | 53.0 (36.0, 65.0) | 0.103 |
| APACHE II score | 21.5 (13.0, 27.0) | 23.0 (15.0, 30.2) | 22.0 (14.0, 29.0) | 0.013 |
| SOFA score | 5.5 (3.2, 7.0) | 6.0 (4.0, 8.0) | 6.0 (4.0, 7.0) | 0.024 |
| Max Serum Creatinine (μmol/l) | 80.5 (68.0, 98.5) | 154.0 (120.5, 279.5) | 99.5 (75.0, 149.8) | < 0.001 |
| KDIGO stage, n (%) |  |  |  | < 0.001 |
| KDIGO1 | 0 (0.0%) | 71 (68.3%) | 71 (28.4%) |  |
| KDIGO2 | 0 (0.0%) | 13 (12.5%) | 13 (5.2%) |  |
| KDIGO3 | 0 (0.0%) | 6 (5.8%) | 6 (2.4%) |  |
| RRT, n (%) | 0 (0.0%) | 14 (13.5%) | 14 (5.6%) | < 0.001 |
| LPV/r, n (%) | 16 (11.0%) | 33 (31.7%) | 49 (19.6%) | < 0.001 |
| Azithromycin, n (%) | 50 (34.2%) | 60 (57.7%) | 110 (44.0%) | < 0.001 |
| Hydroxychloroquine, n (%) | 52 (35.6%) | 60 (57.7%) | 112 (44.8%) | < 0.001 |
| Anakinra, n (%) | 8 (5.5%) | 6 (5.8%) | 14 (5.6%) | 1.000 |
| Dexamethasone, n (%) | 83 (56.8%) | 34 (32.7%) | 117 (46.8%) | < 0.001 |
| NSAID, n (%) | 12 (8.2%) | 17 (16.3%) | 29 (11.6%) | 0.070 |
| Remdesivir, n (%) | 19 (13.0%) | 9 (8.7%) | 28 (11.2%) | 0.315 |
| Steroids, n (%) | 50 (34.2%) | 45 (43.3%) | 95 (38.0%) | 0.186 |
| Antibiotics, n (%) | 137 (93.8%) | 103 (99.0%) | 240 (96.0%) | 0.049 |
| NMBA, n (%) | 90 (61.6%) | 88 (84.6%) | 178 (71.2%) | < 0.001 |
| Noradrenaline, n (%) | 110 (75.3%) | 100 (96.2%) | 210 (84.0%) | < 0.001 |
| Mechanical ventilation, n (%) | 116 (79.5%) | 102 (98.1%) | 218 (87.2%) | < 0.001 |
| Prone positioning, n (%) | 90 (61.6%) | 82 (78.8%) | 172 (68.8%) | 0.004 |
| ECMO, n (%) | 8 (5.5%) | 8 (7.7%) | 16 (6.4%) | 0.602 |
| P/F | 16.4 (11.8, 21.5) | 15.1 (11.9, 20.0) | 16.0 (11.8, 20.7) | 0.333 |
| Tidal volume (mL) | 450.0 (400.0, 480.0) | 460.0 (430.0, 490.0) | 450.0 (420.0, 480.0) | 0.013 |
| Procalcitonin (μg/l) | 0.3 (0.2, 0.8) | 0.7 (0.3, 1.9) | 0.4 (0.2, 1.4) | < 0.001 |
| CRP (mg/l), | 127.0 (76.3, 193.0) | 160.8 (113.0, 216.2) | 138.0 (85.0, 205.0) | 0.018 |
| White blood cells (G/l) | 9.8 (6.3, 13.1) | 9.2 (6.5, 11.2) | 9.5 (6.4, 12.6) | 0.426 |
| Lactate (mmol/l) | 1.2 (0.9, 1.7) | 1.0 (0.8, 1.5) | 1.1 (0.8, 1.7) | 0.034 |
| Bilirubin (μmol/l) | 9.0 (6.0, 13.0) | 10.0 (7.0, 15.0) | 9.0 (6.0, 14.0) | 0.184 |
| **Outcomes** |  |  |  |  |
| ICU mortality, n (%) | 35 (24.0%) | 33 (31.7%) | 68 (27.2%) | 0.196 |
| 28-day mortality, n (%) | 111 (76.6%) | 75 (73.5%) | 186 (75.3%) | 0.654 |
| ICU LOS (d), | 10.5 (6.0, 17.0) | 16.5 (12.0, 23.2) | 13.0 (8.0, 21.0) | < 0.001 |
| Hospital LOS (d) | 23.0 (15.0, 31.0) | 31.5 (23.0, 42.2) | 26.0 (16.0, 36.0) | < 0.001 |
| RRT weaning, n (%) | 0 (0.0%) | 6 (5.8%) | 6 (2.4%) | < 0.001 |
| Mechanical ventilation duration | 12.0 (7.0, 18.0) | 14.5 (10.0, 20.0) | 13.0 (8.0, 19.0) | 0.032 |
| Day-28 free days ventilation | 18.5 (11.0, 23.0) | 14.0 (8.8, 18.0) | 16.0 (10.0, 22.0) | < 0.001 |

### Development of a pipeline of analyses

To identify subgroups of AKI patients, we based our approach on unsupervised clustering. However, unlike in previous studies, we did not apply a clustering algorithm on the raw dataset but rather designed a three-step pipeline of analyses. Firstly, we built a nonlinear statistical model to identify factors significantly associated with AKI development in ICU patients and calculated the importance of each predictor for AKI risk at a single patient level. Secondly, we reduced the dimension of this matrix and applied an unsupervised clustering algorithm in order to define AKI patient subgroups. Thirdly, we compared the clinical outcomes among those clusters of AKI patients (**Figure 1**). These three steps are detailed in the following paragraphs.

**Figure 1.**
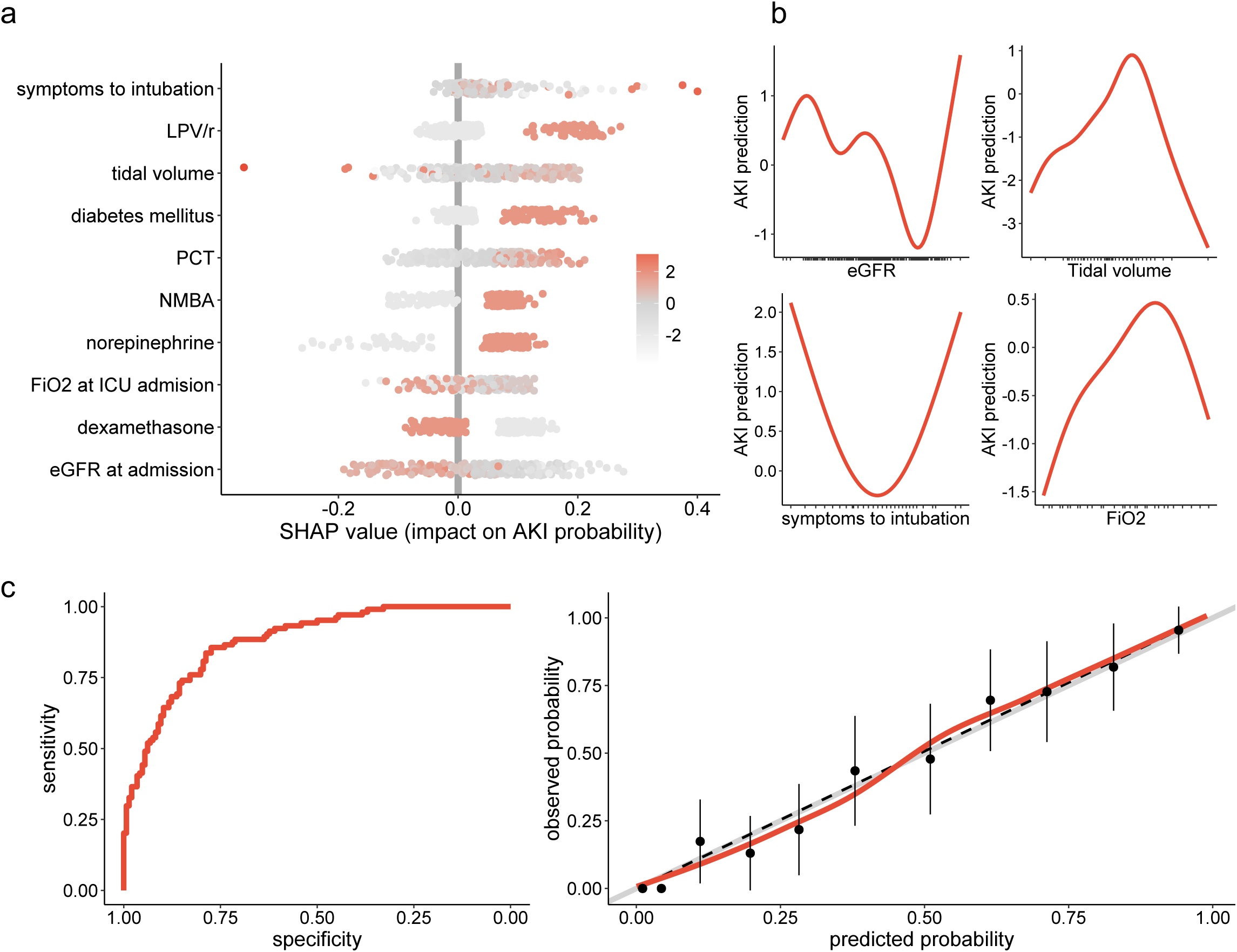
AKI associated factors: a) Shapley Additive Explanation (SHAP) values, where one dot represents the importance of each variable for AKI risk at the single patient level. Positive values reflect an increased risk of AKI while negative values show a negative effect on AKI risk. The sum of the all SHAP values from one patient represent the predicted AKI probability for this patient. Each dot is colour coded according to the patient’s initial value for each considered feature. b) Partial dependence plots, showing the effect of eGFR, tidal volume, time between symptom onset, and intubation as well as FiO2 at ICU admission on the risk of AKI. c) evaluation of the generalized additive model with the receiver operating characteristic curve (left panel) and the calibration plot (right panel) showing sensitivity according to the specificity and the observed versus predicted probabilities respectively. LPV/r Lopinavir/Ritonavir, DM Diabetes Mellitus; PCT Procalcitonin; NMBA Neuromuscular Blocking Agents; DXM Dexamethasone.

### Identification of AKI associated factors

#### Explicative statistical model

We first aimed at identifying factors associated with AKI development in Covid-19 patients admitted to the ICU. We began by preprocessing the data by following three steps. Firstly, numerical variables were centered, scaled and normalized through a Yeo-Johnson transformation, because independent variables were on very different scales and also to enhance variable selection robustness[23]. **Additional File 1** shows the distribution of the numerical variables before and after treatment. Secondly, we imputed missing data using bagged tree imputation [18] to improve accuracy of downstream [24]. Missing data and their distribution for each variable before and after the imputation are presented in **Additional File 2**. Thirdly, we calculated a correlation matrix to identify colinear variables, and removed or merged those with a correlation coefficient above 0.8 (**Additional File 3)**.

Using these pre-processed data, we fitted the relation between AKI and each potential predictor in a univariable fashion, using a logistic regression. As nonlinear relations between AKI and risk factors have often been reported [25–30] we also fitted a logistic regression using natural restricted cubic splines with two degrees of freedom.

We thus performed multivariable analyses using a generalized additive model with a logit link function to allow nonlinear modelling via regression splines. Variables displaying a p-value for univariable association with AKI below 0.2 were primary picked out and feature selection was further achieved using a supervised stepwise approach as previously described [19,20]. The final model identified 10 variables, which were significantly associated with AKI development in the ICU (**Additional Table 1)**: use of LPV/r, NMBA and norepinephrine as well as diabetes mellitus and PCT levels were all positively associated with AKI while administration of dexamethasone was protective. Time between symptom onset and oro-tracheal intubation, eGFR at hospital entrance, tidal volume and FiO2 at ICU admission displayed a nonlinear association with AKI. To estimate the relative contribution of each factor to the predicted probability of AKI, we calculated the Shapley Additive Explanation (SHAP) values. SHAP values represent a feature’s role in changing the model output. In our study, 10 SHAP values were calculated per patient (negative or positive), one for each factor. The sum of each patient SHAP values refers to the predicted AKI probability of this patient. **Figure 1a** displays the SHAP value (x-axis) for each predictor and each patient, while the color of the dot refers to the original value taken by the variable for each patient being considered. Using this strategy, we were able to classify each predictor according to their importance in predicting AKI. Seeing as the relationship between AKI probability and numerical variables were nonlinear, their marginal effect was shown in **Figure 1b**.

Altogether, the final generalized additive model was a discriminant in predicting an AKI ROC curve equal to 0.88 (95% confidence interval [0.84-0.92]), which was well calibrated (p-value of the Hosmer– Lemeshow test equal to 0.97), **Figure 1c**.

#### Sensitivity Analyses

In order to validate the non-linear relationship between the risk of AKI and baseline eGFR, tidal volume, FiO2 at ICU admission and time between symptom onset and oro-tracheal intubation, we first built a second generalized additive model using a local regression by locally estimated scatterplot smoothing curve fitting instead of regression splines. The resulting partial dependence plots (**Additional File 4a)**, showing the marginal effect of each predictor on the risk of AKI, displayed a similar shape to those obtained using regression splines.

Secondly, to ensure the robustness of the variable selection, we applied machine learning (ML) algorithms including native feature selection to predict AKI. We then ran multivariate adaptive regression spline (MARS), multistep adaptative MCPnet, LASSO regression and regularized random forest (RRF) on the entire dataset. A hyperparameter grid was used to tune each model whose performance was iteratively assessed by the area under the ROC curve through a repeated cross-validation procedure. The optimal model was that which maximized the area under the ROC curve. **Additional File 4b** shows the distribution of the out-of-bag area under the ROC curve metric for each predictive model, ranging from 0.73±0.1 to 0.77±0.1 for MCPnet and LASSO models respectively, without significant differences among models. The features selected by each ML algorithm in order of importance in AKI prediction are displayed in **Additional File 4c**. Use of dexamethasone, LPV/r, norepinephrine, eGFR at ICU admission and prior history of diabetes were chosen for every method, while tidal volume, duration between symptom onset and intubation, PCT level, FiO2 at admission and BMI variables were only captured by a nonlinear method (RRF).

Altogether, this sensitivity analysis strengthens both the use of nonlinear fitting between numerical predictors and risk of AKI, as well as the choice of the predictors.

### Identification of AKI phenotypes

#### Clusterisation of AKI patients according to their risk factors pattern

In this second part, we aimed at defining clusters of patients, according to the pattern of risk factors expressed by each patient. To achieve this, we first selected AKI patients and started by reducing the dimension of the matrix of SHAP values previously calculated (104 patients x 10 predictors), using the Uniform Manifold Approximation and Projection (UMAP) method. This preliminary step has been shown to improve downstream clusterisation[31]. This two-dimension projection was finally clusterized by the unsupervised Density-Based Spatial Clustering of Application with Noise (DBSCAN) algorithm. Among the 104 AKI patients, we were able to identify three clusters, each of them expressing a specific pattern of AKI-related factors (**Figure 2a**). For each of them, we characterized their pattern of AKI related factors using the SHAP values. **Figure 2b** shows the predictors in order of importance for each cluster. Cluster 1 was characterized by AKI associated with the use of LPV/r; cluster 2 involves diabetic patients that did not receive dexamethasone; cluster 3 includes patients with low baseline eGFR, long time between symptom onset and orotracheal intubation, high tidal volume and use of norepinephrine. To validate this clusterisation, we defined three subspaces involving two clusters, the cluster of interest and all the others merged. We further applied, on those three subspaces, a support vector machine (SVM) algorithm, and assessed its ability to separate the cluster of interest from the others by a hyperplane. For this purpose, we first randomly split the original cohort into a train and a test dataset, in a 0.8:0.2 ratio. The model was evaluated via a repeated k-fold cross-validation, to find the optimal hyperparameters which maximized the area under the ROC curve. The best model was finally applied on both the train and the test datasets and standard deviations were estimated by bootstrapping. SVM was able to separate each of the three clusters form the others with areas under ROC curves in the test dataset equal to 1.0±0 for each cluster.

**Figure 2.**
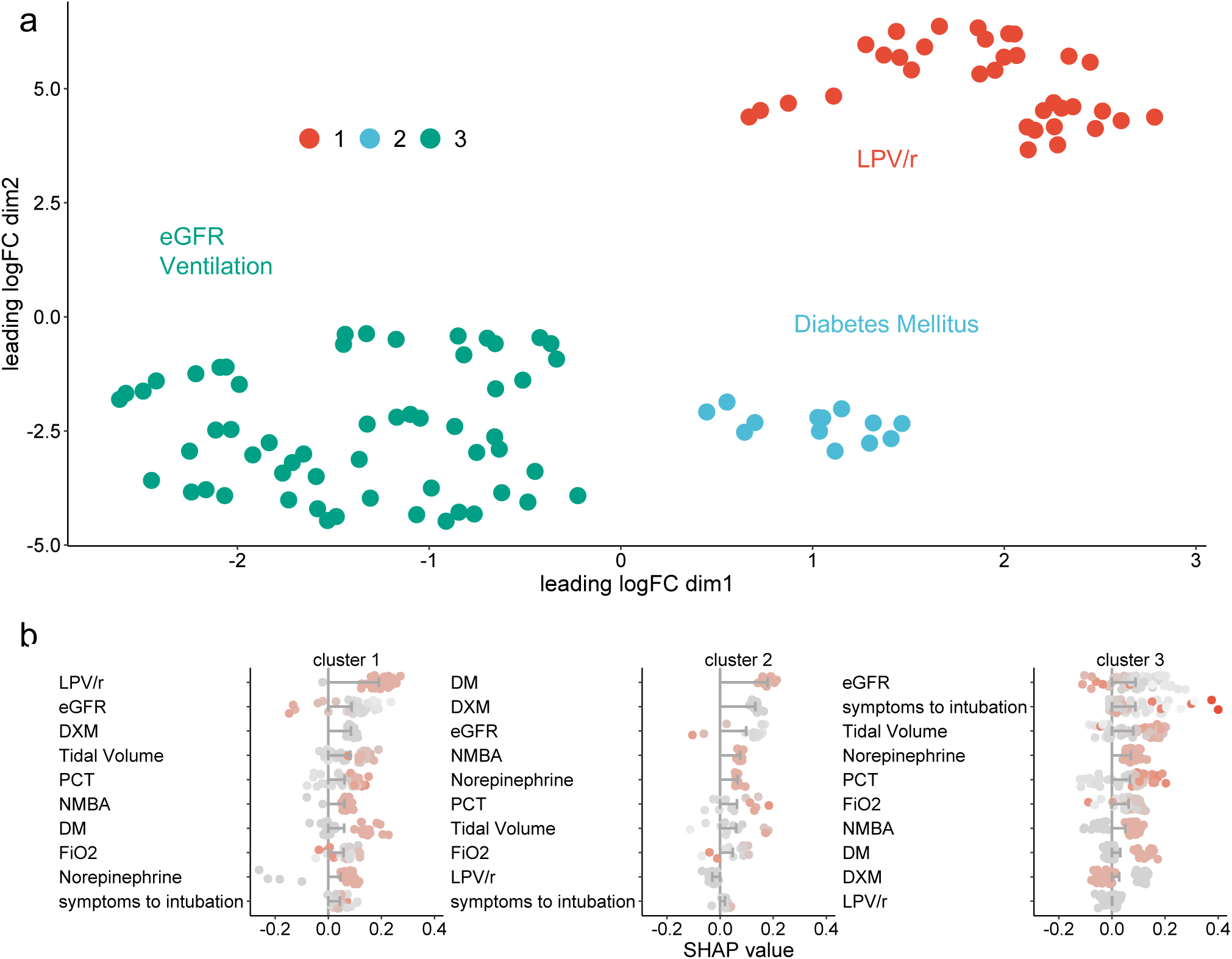
AKI phenotypes’: a) Scatterplot showing the cluster of AKI patients projected on the UMAP, b) Shapley Additive Explanation (SHAP) values for each cluster of AKI patients, sorted by impact on AKI prediction. Bars represent the mean impact of each AKI associated factor for each cluster and dots represent individual patients. LPV/r Lopinavir/Ritonavir, DM Diabetes Mellitus; PCT Procalcitonin; NMBA Neuromuscular Blocking Agents; DXM Dexamethasone.

#### Clinical characteristics of the three AKI phenotypes

Subsequently, we aimed at comparing these three clusters from a clinical perspective. To achieve this objective, we used naïve Bayes classifier in the five subspaces described in the previous paragraph to calculate the a posteriori probability of clinical endpoints, that were bootstrapped 2000 times. As outcomes, we investigated AKI severity and recovery, ICU mortality and relative time spent into one of the following metabolic profile: baseline (normal glucose/normal lactate levels), isolated hyperglycaemia (high glucose but normal lactate levels), isolated hypoglycaemia (low glucose but normal lactate levels), stress response (high glucose/high lactate) and impaired metabolism (low to normal glucose level with high lactate level), as previously described [21]. These five metabolic profiles are shown in **Additional File 5**.

#### Renal endpoints

Patients from clusters 1 and 2 developed more severe AKI than patients from cluster 3 (34% [21-47] versus 7% [0-12] of KDIGO3 AKI, p=0.02) and more frequently received RRT (25% [13-38] versus 4% [0-7], p<0.001). They also showed lower renal function recovery rates (51% [35-68] versus 82% [71-97] of the patients fully recover their renal function while 14% [3-24] versus 0% [0-0] were not weaned from the RRT at ICU discharge p=0.006 and 0.008 respectively, **Figure 3a,b**). However, midterm recovery of renal function, defined as a 50% decrease in serum creatinine from the maximum levels reached during ICU stay, was better for patients in cluster 1, compared to the two other clusters (Hazard ratio equal to 2.0 [1.0-4.1], p=0.046, **Figure 3c**).

**Figure 3.**
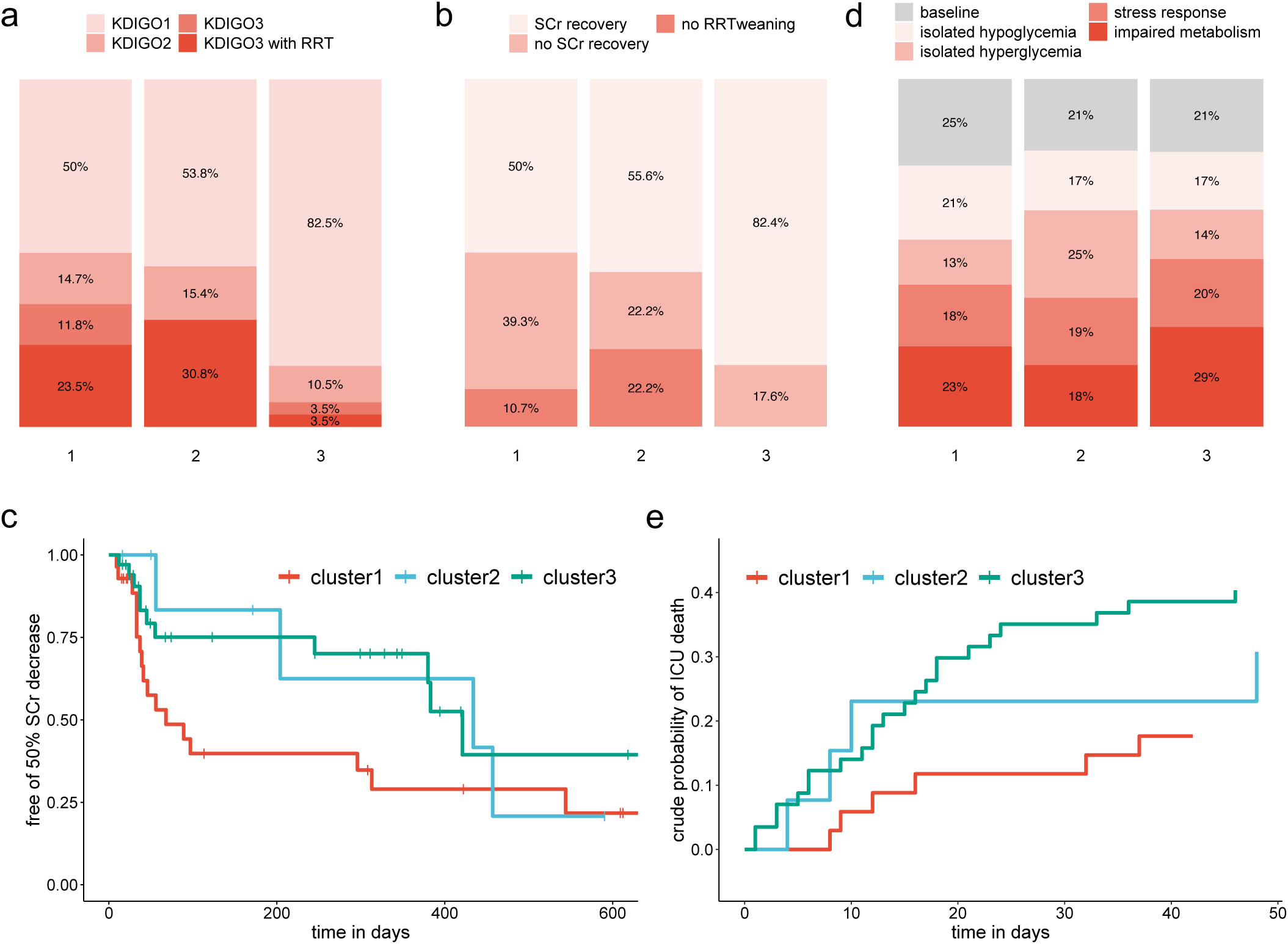
Clinical outcomes of each cluster: a) repartition of AKI severity among clusters, according to the KDIGO criteria b) repartition of renal recovery at ICU discharge, c) survival curve showing the the proportion of patients who did not experience serum creatinine reduction above 50% from the maximal level reached during ICU stay, over time and among clusters, d) relative time spent in each metabolic pattern according to clusters of AKI patients and e) cumulative incidence curve of the ICU mortality, stratified on clusters of AKI patients and c) eGFR estimated Glomerular Filtration Rate; RRT Renal Replacement Therapy .p-value<0.1; *p-value<0.05; **p-value<0.001; ***p-value<0.0001

Patients from clusters 1 and 2 also showed a distinct metabolic profile compared to cluster 3, expressing the impaired metabolism profile at a lower rate (22% [19-24] versus 29% [23-33] p=0.005, **Figure 3d**). Among clusters 1 and 2, the expression of isolated hyperglycaemia was also higher in cluster 2 (13% [10-16] versus 25% [11-35], p=0.009, **Figure 3d**).

Finally, mortality varied between the 3 clusters (18% [3-29], 30% [8-54] and 40% [28-52] respectively, **Figure 3e**).

Altogether, this analytic procedure allowed us to identify 3 clusters of AKI patients, each of them expressing a specific pattern of factors associated with AKI. These patients also displayed different clinical characteristics, including different AKI severity, mortality and recovery.

## Discussion

The current definition of AKI is limited as it provides no information on AKI etiology, prognosis, molecular pathways, or responses to treatment[32]. Here we identified phenotypes of AKI patients based on their pattern of AKI associated factors, with distinct characteristics and outcomes.

We first identified factors associated with AKI development. When considering Covid19 specific therapy, we found LPV/r and dexamethasone to be respectively positively and negatively related to AKI development, in accordance with other groups[33–37]. We also reported well described AKI risk factors, such as diabetes mellitus, use of norepinephrine and baseline eGFR[38,39]. Interestingly, the relation between AKI risk and the baseline eGFR was nonlinear, where higher eGFRs were associated with a reduced probability of AKI up to a threshold beyond which the risk increased. We also found a linear relation between AKI development and procalcitonin (PCT) levels at ICU admission. Several studies have reported an increased PCT level in patients with reduced GFR[40–42] suggesting that PCT may be partially removed via the kidneys. More recently, PCT has also been shown to predict AKI[43–45]. Finally, we identified four factors related to mechanical ventilation, the time between symptoms onset and intubation, the tidal volume, the FiO2 at ICU admission and the use of NMBA. While very few studies have assessed the impact of these factors on AKI development, several of them confirm the association between mechanical ventilation requirement and AKI occurrence in Covid-19 patients[46,47].

In our cohort of AKI Covid19 patients, our pipeline was able to identify three clusters of patients. At the renal level, while all patients met the criteria for AKI, each cluster display a distinct phenotype in terms of severity, ICU recovery and mid-term recovery. In particular, cluster 1 involving patients receiving LPV/r was characterized by severe AKI with 24% of patients requiring renal replacement therapy. At ICU discharge, only 50% of them had recovered their renal function but, paradoxically, they experienced better mid-term recovery and the lowest ICU mortality rate. At the metabolic level, we found that patients from cluster 3 had the highest ICU mortality, and were mostly in the impaired metabolism profile, as previously shown by our group[21,22]. Similarly, diabetic patients in cluster 2 displayed a higher rate of the isolated hyperglycemia pattern. Altogether, these three phenotypes may reflect distinct pathophysiological mechanisms of AKI development.

Beyond these results, this study introduces a pipeline of analyses, which are able to phenotype AKI patients according to their pattern of risk factors, with several innovative features. Firstly, while most of the studies identified AKI risk factors through logistic regression[47,48], we used a generalized additive model with regression splines to capture nonlinear associations between AKI and potential risk factors. This method allowed us to identify factors that would have remained otherwise unnoticed with the traditional approach (i.e. baseline eGFR, tidal volume, time between symptoms onset and intubation and FiO2 at ICU admission). Furthermore, the absolute importance of each risk factor in estimating the probability of AKI we calculated for each patient. We thus obtained a pattern of risk factors for each patient that may reflect a specific pathophysiological mechanism. Existing studies on AKI phenotyping have either used supervised clustering, mostly on clinical traits[13,14], or unsupervised clustering based on recorded clinical or biological data[15–17]. Finally, we did not apply the clustering algorithm on the raw dataset as did other groups[15–17]., but rather on a dimensionally reduced space; a strategy that has been shown to improve the clustering performance[31].

Our study has some limitations. Firstly, the study was single-centred which limits the extent of our results. Secondly, being a retrospective study, procedures and therapeutic strategies may have changed during the study period. Nonetheless, we feel we have provided a generalizable pipeline that may be applied to various datasets to identify patients with different outcomes and therapeutic sensitivity.

## Conclusion

We have developed a new pipeline of analyses in order to identify the phenotype of AKI patients based on their pattern of AKI risk factors when applying this method to a Covid19 ICU patient dataset. We identified 3 patient subgroups with distinct renal features and outcomes that may be related to specific pathophysiological mechanisms.

## Data Availability

The datasets used and/or analysed during the current study are available from the corresponding author on reasonable request.

## Declarations

### Ethics approval and consent to participate

The study was approved by the local ethical committee for human studies of Geneva, Switzerland (CCER 2020-00917, Commission Cantonale d’Ethique de la Recherche) and performed according to the Declaration of Helsinki principles.

### Consent for publication

Not applicable

### Authors’ contributions

Conceptualization, D.L.; methodology, D.L. and G.C.; validation, D.L., G.C. and J.P..; formal analysis, D.L., G.C.; data curation, D.L., F.S., E.M., C.L.; writing—original draft preparation, D.L., F.S., E.M..; writing—review and editing, D.L., G.C., C.L., S.S., J.P.; supervision, D.L. All authors have read and agreed to the published version of the manuscript.

## Acknowledgements

Not applicable

## Figures legend

**Additional File 1.**
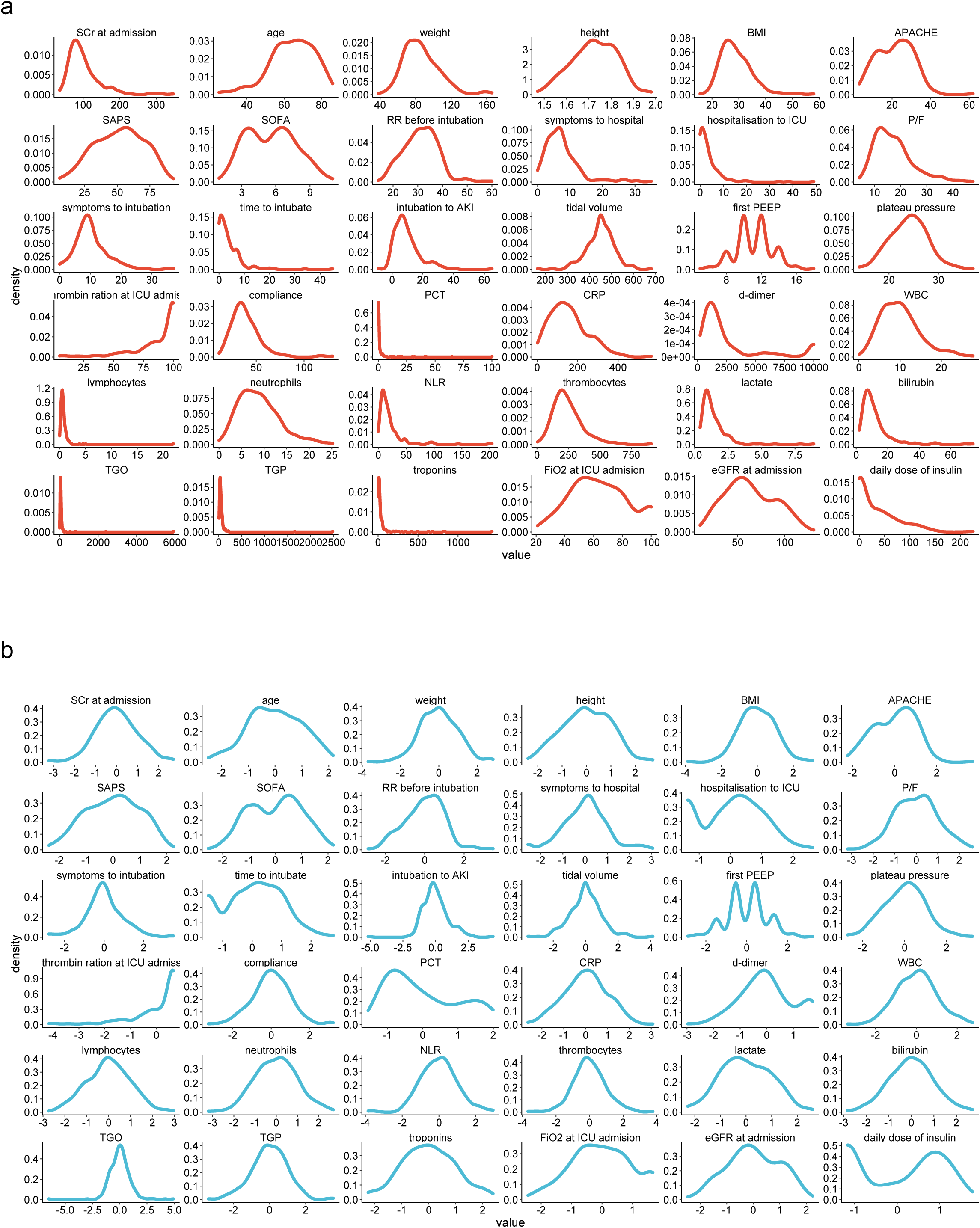
data transformation: distribution of the numerical variables before (a) and after (b) scaling, centering and Yeo-Johnson transformation. SCr Serum Creatinine; BMI Body Mass Index; RR Respiratory Rate; P/F PaO2/FiO2 ratio; PCT Procalcitonin; CRP C-Reactive Protein; WBC White Blood Cells; NLR Neutrophils to Lymphocytes Ratio; PP Prone Positioning; NMBA Neuro Muscular Blocking Agents.

**Additional File 2.**
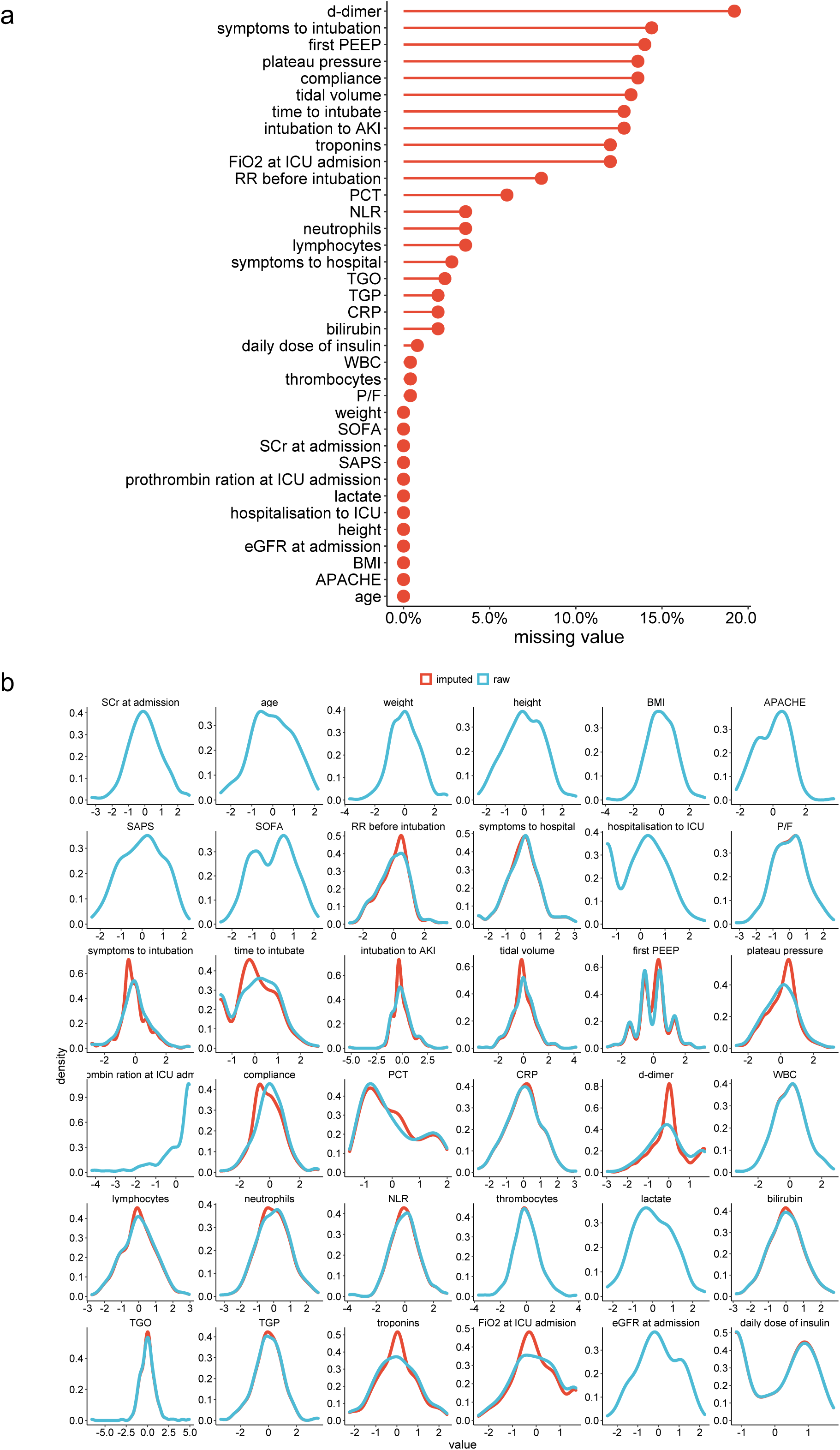
handling of missing value: a) missing value rate for each recorded variable and b) distribution of the variable before (blue) and after (red) data imputation. SCr Serum Creatinine; BMI Body Mass Index; RR Respiratory Rate; P/F PaO2/FiO2 ratio; PCT Procalcitonin; CRP C-Reactive Protein; WBC White Blood Cells; NLR Neutrophils to Lymphocytes Ratio; PP Prone Positioning; NMBA Neuro Muscular Blocking Agents.

**Additional File 3.**
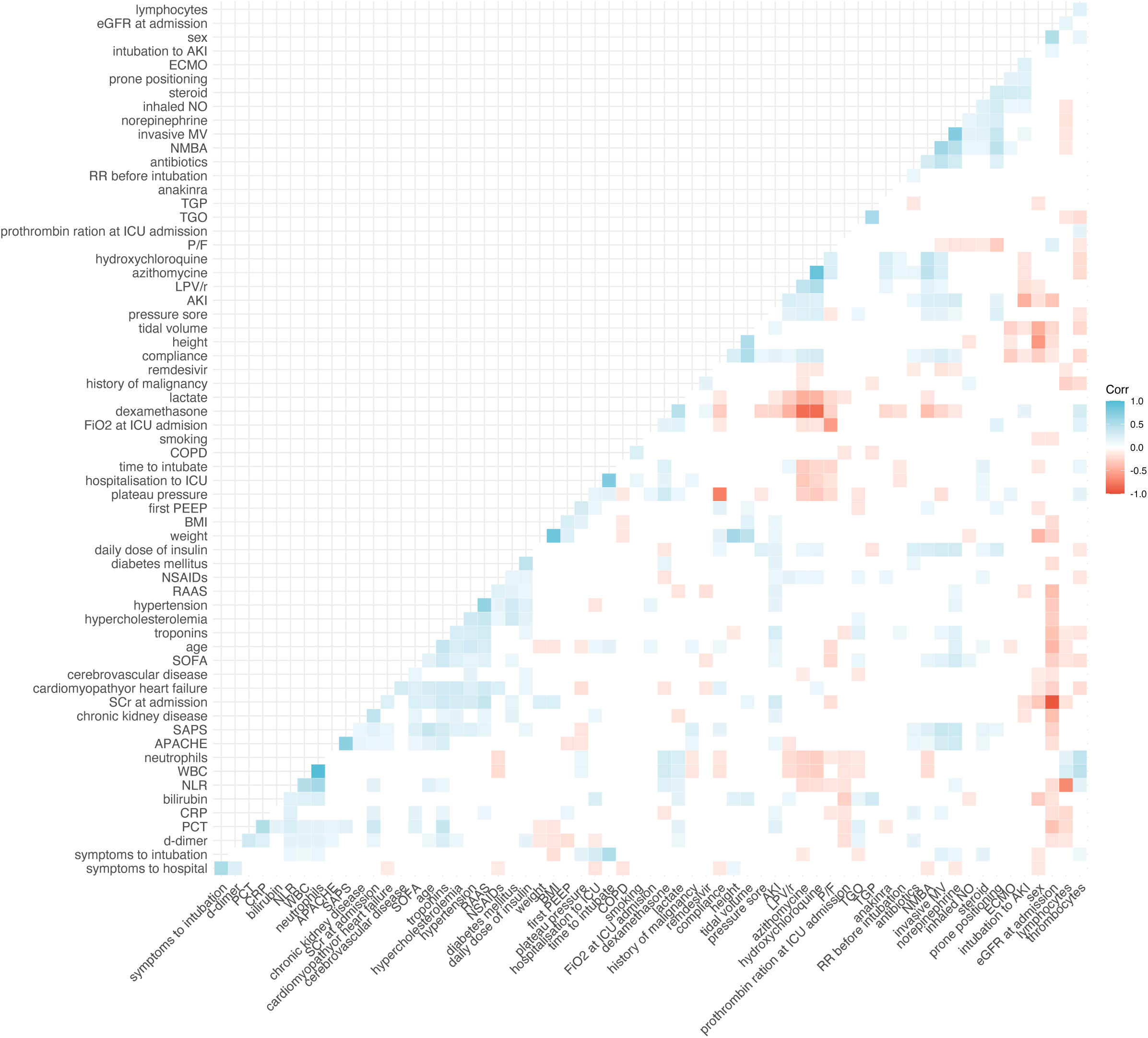
correlated variables: correlation matrix showing the correlation between each recorded variable. Blank cases mean non-significant correlation (p-value>0.05). BMI Body Mass Index; NO Nitric Oxide; NMBA Neuro Muscular Blocking Agents; WBC White Blood Cells; NLR Neutrophils to Lymphocytes Ratio; PCT Procalcitonin; CRP C-Reactive Protein; eGFR estimated Glomerular Filtration Rate; P/F PaO2/FiO2 ratio; SCr Serum Creatinine; RAAS Renin Angiotensin Aldosterone System blockers; NSAIDs Non-Steroidal Anti-inflammatory Drugs; COPD Chronic Obstructive Pulmonary Disease; LPV/r Lopinavir/Ritonavir, MV Mechanical Ventilation;

**Additional File 4.**
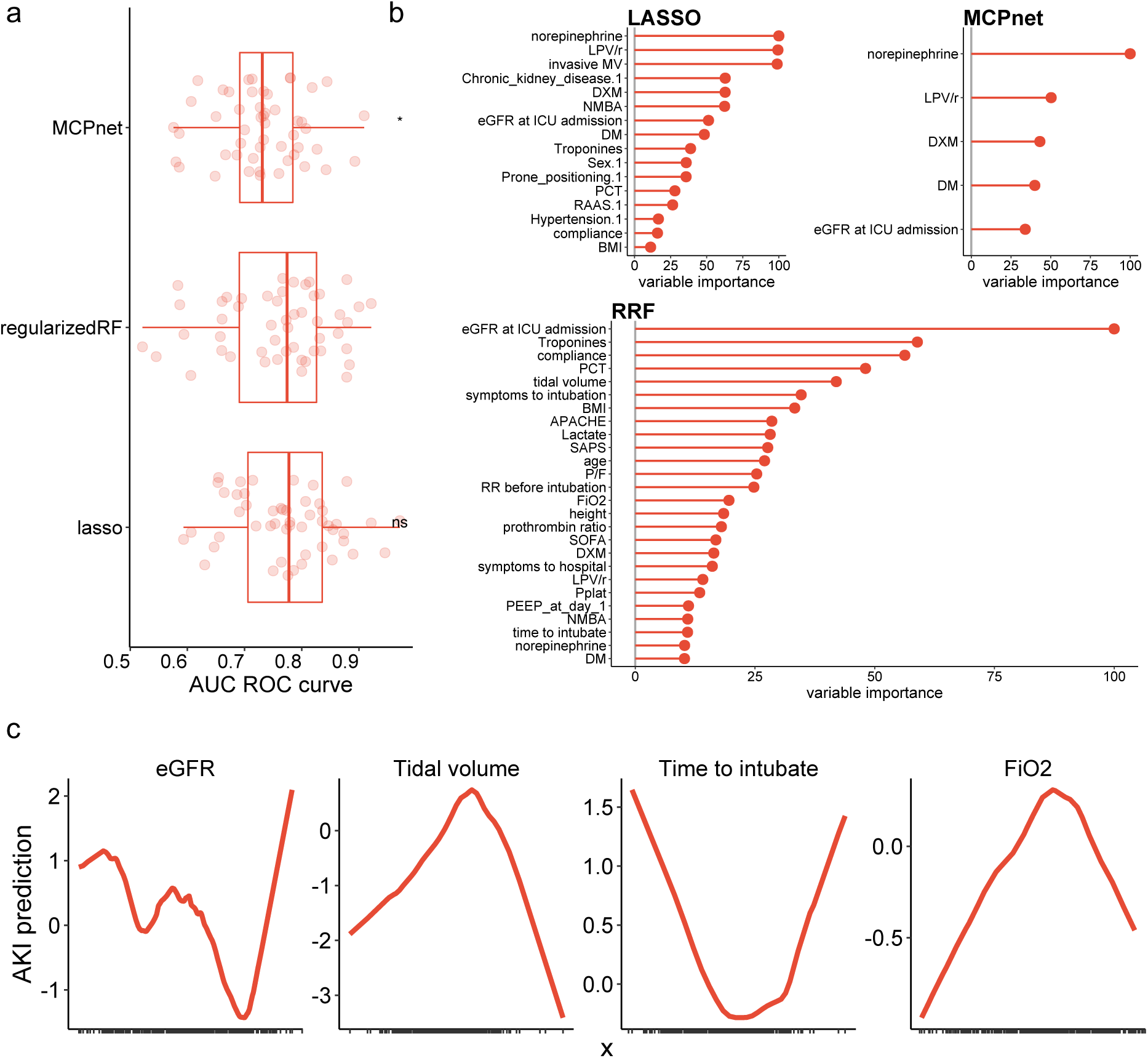
model validation: a) boxplot showing the distribution of the area under the receiver operating characteristic curve for each machine learning algorithm trained to predict AKI, b) importance of the selected variables for AKI prediction for each machine learning method and c) partial dependence plots, showing the effect of BMI, SAPS score and duration from hospitalization to intubation on the risk of AKI, extracted from the generalized additive model with LOESS fitting. LPV/r Lopinavir/Ritonavir; DXM Dexamethasone; eGFR estimated Glomerular Filtration Rate; DM Diabetes Mellitus; BMI Body Mass index; P/F PaO2/FiO2 ratio; RR Respiratory Rate.

**Additional File 5.**
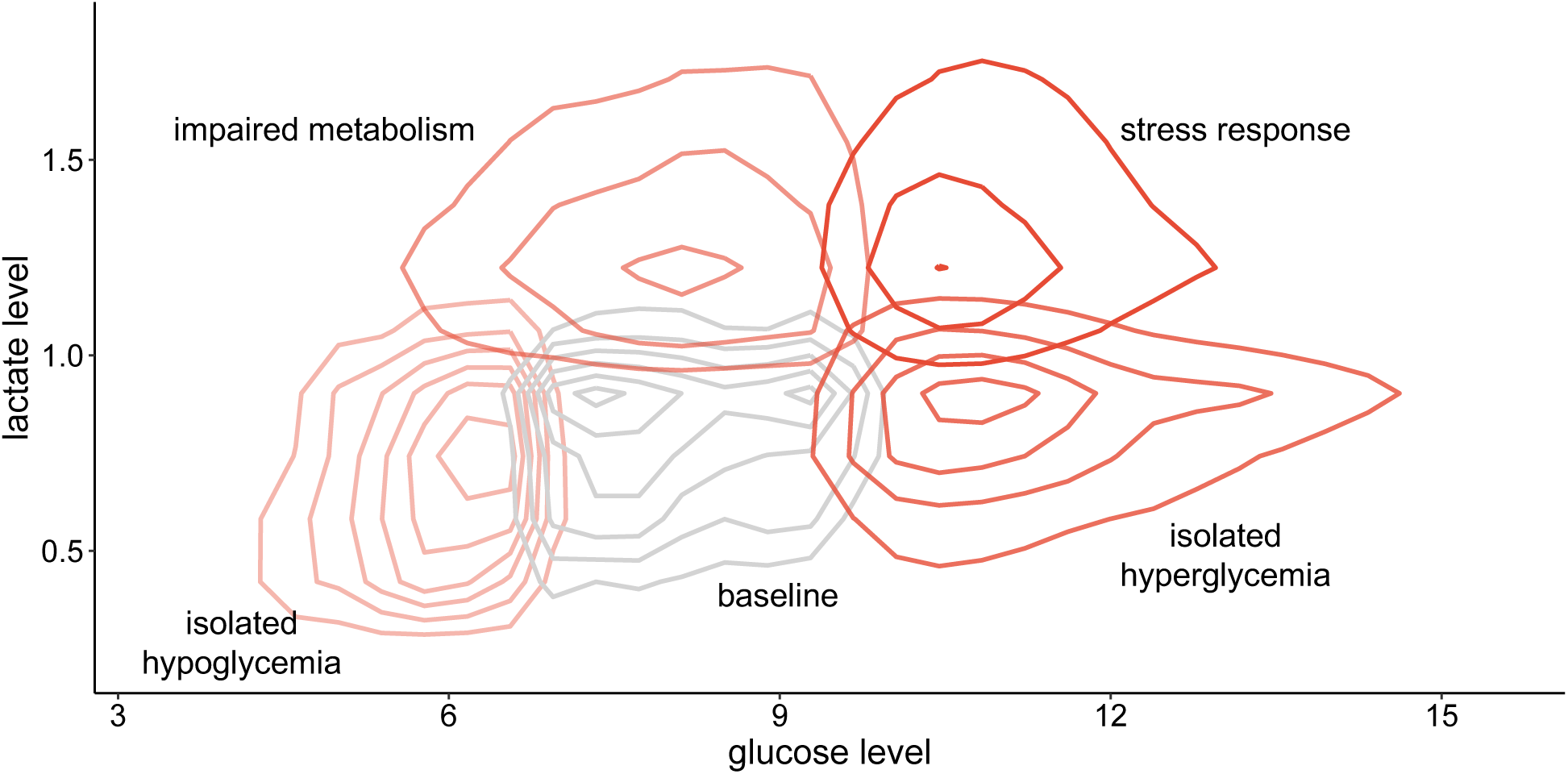
definition of the 5 metabolic patterns: density plot showing lactate and glucose levels in mmol/L according to the five metabolic patterns defined.

**Additional Table 1:** variables selected in the final generalized additive mode for AKI prediction with corresponding pvalue. LPV/r Lopinavir/Ritonavir; PCT Procalcitonin; NMBA Neuromuscular Blocking Agent; eGFR estimated Glomerular Filtration Rate

| variable | pvalue |
| --- | --- |
| LPV/r | 0.009 |
| PCT | 0.006 |
| norepinephrine | 0.001 |
| dexamethasone | 0.009 |
| NMBA | 0.049 |
| diabetes mellitus | <0.001 |
| eGFR at Hospital entrance | 0.012 |
| Tidal volume | 0.002 |
| Time between symptoms onset and intubation | 0.004 |
| FiO2 at ICU admission | 0.040 |

## References

1. Hoste EAJ, Bagshaw SM, Bellomo R, Cely CM, Colman R, Cruz DN, et al. Epidemiology of acute kidney injury in critically ill patients: the multinational AKI-EPI study. Intensive Care Medicine. 2015;41:1411–23.

2. Nisula S, Kaukonen K-M, Vaara ST, Korhonen A-M, Poukkanen M, Karlsson S, et al. Incidence, risk factors and 90-day mortality of patients with acute kidney injury in Finnish intensive care units: the FINNAKI study. Intensive Care Med. 2013;39:420–8.

3. Gaudry S, Hajage D, Schortgen F, Martin-Lefevre L, Pons B, Boulet E, et al. Initiation Strategies for Renal-Replacement Therapy in the Intensive Care Unit. New England Journal of Medicine. 2016;375:122–33.

4. Gaudry S, Hajage D, Martin-Lefevre L, Lebbah S, Louis G, Moschietto S, et al. Comparison of two delayed strategies for renal replacement therapy initiation for severe acute kidney injury (AKIKI 2): a multicentre, open-label, randomised, controlled trial. The Lancet. Elsevier; 2021;397:1293–300.

5. The STARRT-AKI Investigators. Timing of Initiation of Renal-Replacement Therapy in Acute Kidney Injury. New England Journal of Medicine. 2020;383:240–51.

6. Barbar SD, Clere-Jehl R, Bourredjem A, Hernu R, Montini F, Bruyère R, et al. Timing of Renal-Replacement Therapy in Patients with Acute Kidney Injury and Sepsis. New England Journal of Medicine. 2018;379:1431–42.

7. Kellum JA, Lameire N, Aspelin P, Barsoum RS, Burdmann EA, Goldstein SL, et al. KDIGO Clinical Practice Guideline for Acute Kidney Injury 2012. Kidney International Supplements. 2012;2:1–138.

8. Castela Forte J, Perner A, van der Horst ICC. The use of clustering algorithms in critical care research to unravel patient heterogeneity. Intensive Care Med. 2019;45:1025–8.

9. Endre ZH, Mehta RL. Identification of acute kidney injury subphenotypes. Curr Opin Crit Care. 2020;26:519–24.

10. Fereshtehnejad S-M, Zeighami Y, Dagher A, Postuma RB. Clinical criteria for subtyping Parkinson’s disease: biomarkers and longitudinal progression. Brain. Oxford University Press; 2017;140:1959–76.

11. Zhang X, Chou J, Liang J, Xiao C, Zhao Y, Sarva H, et al. Data-driven subtyping of Parkinson’s disease using longitudinal clinical records: a cohort study. Scientific reports. Nature Publishing Group; 2019;9:1–12.

12. Jannot A-S, Burgun A, Thervet E, Pallet N. The Diagnosis-Wide Landscape of Hospital-Acquired AKI. Clin J Am Soc Nephrol. 2017;12:874–84.

13. Bhatraju PK, Mukherjee P, Robinson-Cohen C, O’Keefe GE, Frank AJ, Christie JD, et al. Acute kidney injury subphenotypes based on creatinine trajectory identifies patients at increased risk of death. Crit Care. 2016;20:372.

14. Kellum JA, Sileanu FE, Bihorac A, Hoste EAJ, Chawla LS. Recovery after Acute Kidney Injury. American Journal of Respiratory and Critical Care Medicine. 2017;195:784–91.

15. Wiersema R, Jukarainen S, Vaara ST, Poukkanen M, Lakkisto P, Wong H, et al. Two subphenotypes of septic acute kidney injury are associated with different 90-day mortality and renal recovery. Crit Care. 2020;24:150.

16. Bhatraju PK, Zelnick LR, Herting J, Katz R, Mikacenic C, Kosamo S, et al. Identification of Acute Kidney Injury Subphenotypes with Differing Molecular Signatures and Responses to Vasopressin Therapy. Am J Respir Crit Care Med. 2019;199:863–72.

17. Chaudhary K, Vaid A, Duffy Á, Paranjpe I, Jaladanki S, Paranjpe M, et al. Utilization of Deep Learning for Subphenotype Identification in Sepsis-Associated Acute Kidney Injury. Clin J Am Soc Nephrol. 2020;15:1557–65.

18. Jäger S, Allhorn A, Bießmann F. A Benchmark for Data Imputation Methods. Frontiers in Big Data. 2021;4:48.

19. Bursac Z, Gauss CH, Williams DK, Hosmer DW. Purposeful selection of variables in logistic regression. Source Code for Biology and Medicine. 2008;3:17.

20. Legouis D, Jamme M, Galichon P, Provenchère S, Boutten A, Buklas D, et al. Development of a practical prediction score for chronic kidney disease after cardiac surgery. Br J Anaesth. 2018;121:1025–33.

21. Legouis D, Ricksten S-E, Faivre A, Verissimo T, Gariani K, Verney C, et al. Altered proximal tubular cell glucose metabolism during acute kidney injury is associated with mortality. Nat Metab. 2020;2:732–43.

22. Verissimo T, Faivre A, Rinaldi A, Lindenmeyer M, Delitsikou V, Veyrat-Durebex C, et al. Decreased Renal Gluconeogenesis Is a Hallmark of Chronic Kidney Disease. J Am Soc Nephrol. 2022;ASN.2021050680.

23. Peterson R A. Finding Optimal Normalizing Transformations via bestNormalize. The R Journal. 2021;13:310.

24. Jaeger BC, Cantor R, Sthanam V, Xie R, Kirklin JK, Rudraraju R. Improving Outcome Predictions for Patients Receiving Mechanical Circulatory Support by Optimizing Imputation of Missing Values. Circ Cardiovasc Qual Outcomes. 2021;14:e007071.

25. Han SS, Baek SH, Ahn SY, Chin HJ, Na KY, Chae D-W, et al. Anemia Is a Risk Factor for Acute Kidney Injury and Long-Term Mortality in Critically Ill Patients. The Tohoku Journal of Experimental Medicine. 2015;237:287–95.

26. Adhikari L, Ozrazgat-Baslanti T, Ruppert M, Madushani RWMA, Paliwal S, Hashemighouchani H, et al. Improved predictive models for acute kidney injury with IDEA: Intraoperative Data Embedded Analytics. PLoS One. 2019;14:e0214904.

27. Huang C, Li S-X, Mahajan S, Testani JM, Wilson FP, Mena CI, et al. Development and Validation of a Model for Predicting the Risk of Acute Kidney Injury Associated With Contrast Volume Levels During Percutaneous Coronary Intervention. JAMA Network Open. 2019;2:e1916021.

28. Zhou J, Lyu L, Zhu L, Liang Y, Dong H, Chu H. Association of overweight with postoperative acute kidney injury among patients receiving orthotopic liver transplantation: an observational cohort study. BMC Nephrol. 2020;21:223.

29. Thongprayoon C, Cheungpasitporn W, Chewcharat A, Mao MA, Bathini T, Vallabhajosyula S, et al. Impact of admission serum ionized calcium levels on risk of acute kidney injury in hospitalized patients. Sci Rep. 2020;10:12316.

30. Cheng Y, Zhang Y, Tu B, Qin Y, Cheng X, Qi R, et al. Association Between Base Excess and Mortality Among Patients in ICU With Acute Kidney Injury. Frontiers in Medicine. 2021;8:2436.

31. Allaoui M, Kherfi ML, Cheriet A. Considerably Improving Clustering Algorithms Using UMAP Dimensionality Reduction Technique: A Comparative Study. In: El Moataz A, Mammass D, Mansouri A, Nouboud F, editors. Image and Signal Processing. Cham: Springer International Publishing; 2020. p. 317–25.

32. Waikar SS, Betensky RA, Emerson SC, Bonventre JV. Imperfect gold standards for kidney injury biomarker evaluation. J Am Soc Nephrol. 2012;23:13–21.

33. Mousavi Movahed SM, Akhavizadegan H, Dolatkhani F, Nejadghaderi SA, Aghajani F, Faghir Gangi M, et al. Different incidences of acute kidney injury (AKI) and outcomes in COVID-19 patients with and without non-azithromycin antibiotics: A retrospective study. Journal of Medical Virology. 2021;93:4411–9.

34. Binois Y, Hachad H, Salem J-E, Charpentier J, Lebrun-Vignes B, Pène F, et al. Acute Kidney Injury Associated With Lopinavir/Ritonavir Combined Therapy in Patients With COVID-19. Kidney International Reports. 2020;5:1787–90.

35. Schneider J, Jaenigen B, Wagner D, Rieg S, Hornuss D, Biever PM, et al. Therapy with lopinavir/ritonavir and hydroxychloroquine is associated with acute kidney injury in COVID-19 patients. PLoS One. 2021;16:e0249760.

36. Grimaldi D, Aissaoui N, Blonz G, Carbutti G, Courcelle R, Gaudry S, et al. Characteristics and outcomes of acute respiratory distress syndrome related to COVID-19 in Belgian and French intensive care units according to antiviral strategies: the COVADIS multicentre observational study. Ann Intensive Care. 2020;10:131.

37. Orieux A, Khan P, Prevel R, Gruson D, Rubin S, Boyer A. Impact of dexamethasone use to prevent from severe COVID-19-induced acute kidney injury. Critical Care. 2021;25:249.

38. Sanchez-Russo L, Billah M, Chancay J, Hindi J, Cravedi P. COVID-19 and the Kidney: A Worrisome Scenario of Acute and Chronic Consequences. J Clin Med. 2021;10:900.

39. Smith LE, Smith DK, Blume JD, Siew ED, Billings FT. Latent variable modeling improves AKI risk factor identification and AKI prediction compared to traditional methods. BMC Nephrology. 2017;18:55.

40. Nakamura Y, Murai A, Mizunuma M, Ohta D, Kawano Y, Matsumoto N, et al. Potential use of procalcitonin as biomarker for bacterial sepsis in patients with or without acute kidney injury. J Infect Chemother. 2015;21:257–63.

41. Steinbach G, Bölke E, Grünert A, Orth K, Störck M. Procalcitonin in patients with acute and chronic renal insufficiency. Wien Klin Wochenschr. 2004;116:849–53.

42. Park JH, Kim DH, Jang HR, Kim M-J, Jung S-H, Lee JE, et al. Clinical relevance of procalcitonin and C-reactive protein as infection markers in renal impairment: a cross-sectional study. Critical Care. 2014;18:640.

43. Chun K, Chung W, Kim AJ, Kim H, Ro H, Chang JH, et al. Association between acute kidney injury and serum procalcitonin levels and their diagnostic usefulness in critically ill patients. Sci Rep. Nature Publishing Group; 2019;9:4777.

44. Hu Q, Zhang Y, Xu H, Zhu L, Chen L, Hao C. Association between admission serum procalcitonin and the occurrence of acute kidney injury in patients with septic shock: A retrospective cohort study. Science Progress. SAGE Publications Ltd; 2021;104:00368504211043768.

45. Kan W-C, Huang Y-T, Wu V-C, Shiao C-C. Predictive Ability of Procalcitonin for Acute Kidney Injury: A Narrative Review Focusing on the Interference of Infection. Int J Mol Sci. 2021;22:6903.

46. Cai X, Wu G, Zhang J, Yang L. Risk Factors for Acute Kidney Injury in Adult Patients With COVID-19: A Systematic Review and Meta-Analysis. Front Med (Lausanne). 2021;8:719472.

47. Hirsch JS, Ng JH, Ross DW, Sharma P, Shah HH, Barnett RL, et al. Acute kidney injury in patients hospitalized with COVID-19. Kidney Int. 2020;98:209–18.

48. de Almeida DC, Franco M do CP, Dos Santos DRP, Santos MC, Maltoni IS, Mascotte F, et al. Acute kidney injury: Incidence, risk factors, and outcomes in severe COVID-19 patients. PLoS One. 2021;16:e0251048.

